# PRACTICE OF EARLY BREASTFEEDING INITIATION ON PRIMIGRAVIDA WITH A CULTURAL PERSPECTIVE: A SYSTEMATIC REVIEW

**DOI:** 10.1101/2022.01.15.22269367

**Authors:** Ni Wayan Dewi Tarini, Moses Glorino Rumambo Pandin

## Abstract

One of the health aspects in the development process is the development of human resource potential (HR). What plays a role in this potential of human resources is good nutritional status from an early age. In addition to these factors, the quality of human resource formation is also influenced by the environment that shapes human character. namely the socio-cultural environment (Soekirman, 2005). One of the cultural interactions that affect nutritional status as a condition for the formation of quality human resources is the practice of breastfeeding and the practice of early breastfeeding initiation (IMD). There have been many previous studies that have written that there are still many mothers who do not practice IMD, where one of the reasons mentioned is the belief that the first milk that comes out is dirty and unhealthy, so it is not suitable to be given to babies (Roesli, 2008). The purpose of this literature study is to determine the influence of culture in the practice of early breastfeeding initiation.

The articles used in the literature review were obtained through databases of international journal providers such as Proquest, Google Scholar and Science Direct. The journal was taken from 2019 to 2021. Researchers used keywords, namely transcultural nursing, breastfeeding (according to MESH/Medical Subject Heading) and 20 articles were analyzed using PRISMA diagrams. The analysis found that most mothers did not practice early breastfeeding because of certain beliefs about early breastfeeding, such as early breastfeeding was not as good as exclusive breastfeeding, dirty early breast milk, unhealthy early breastfeeding given to babies and the habit of mothers who immediately gave additional food immediately. after the baby is born.

## INTRODUCTION

The health aspect in the development process one of them is the development of human resource potential. What plays a role in the potential of human resources is good nutritional status from an early age. In addition to these factors, the quality of human resource formation is also influenced by the environment that shapes human character. Socio-cultural environment (Soekirman, 2005). This environment can help shape the interactions, communication, values, norms and cultural ethics that are long established that are passed down through generations. One of the cultural interactions that affect nutritional status as a condition of the establishment of human resource quality is the practice of breastfeeding and the practice of early breastfeeding initiation (EBI). Many previous studies have written that there are still many mothers who do not practice EBI, which is one of the causes mentioned is the belief that the milk that first comes out is unhealthy for the baby so it is not worth giving to the baby (Roesli, 2008).

EBI coverage in Buleleng Regency in 2020 only reached 32% even though the target in Indonesia is 80%. This coverage is still very far below the target in Indonesia, so efforts to improve EBI practices are the focus of local health policies that must be considered by every childbirth helper (Dinkes Bali Province, 2020).

Culture and tradition are inseparable in human life. According to Leininger (2002) humans tend to maintain their habits wherever they are. Leininger divided the socio-cultural dimensions of seven factors: 1) Technological factors; 2) Religious factors and philosophy of life; 3) Social factors and family attachments; 4) Cultural values and way of life; 5) Applicable policy and regulatory factors; 6) Economic factors; and 7) Educational factors. These factors according to Leininger can affect health.

In the practice of EBI, EBI behavior is strongly influenced by the hereditary tradition inherited by the mother from her family, each mother tends to apply different EBI behaviors. So in this condition, the behavior of EBI carried out by the mother can be said to be a cultural practice because it shows different behaviors. Some research reveals that cultural practices that occur in the community can inhibit the practice of EBI, but there are also those who support the practice of EBI. Like research conducted by Mayangsari, R (2019) who wrote IMD can not be separated from the prevailing cultural influence in society. In Kendari City there are still mothers who believe and believe that colostrum is dirty breast milk and giving honey at an early age is good for the health of newborns. Some people think that formula milk is better than breast milk. Ningsih Research, RR (2021) writes that 49,5% of mothers in Posyandu work area of Samarinda New Hope Health Center, the practice of early breastfeeding initiation carried out is strongly influenced by the local culture, where breast milk that comes out first must be discarded because it is dirty breast milk. While Wagrner, S (2019) writes that mothers who have experience doing EBI practice will affect the duration of subsequent breastfeeding.

Traditions and beliefs develop for generations which will cause people’s behavior to lead to the traditions that have been believed. Myths that develop in society in general can affect the practice of EBI, such as the myth that colostrum is stale milk, early delivery of solid food/ honey will be good for the growth and development of children, parenting patterns that develop in the community and supported by low maternal and family knowledge will cause mothers to practice EBI such as beliefs or traditions that are believed to be true for generations. Socio-cultural beliefs come from what was seen before. Trust that has been formed will be the basis of a person’s knowledge and behavior..

## METHOD

Articles used in literature reviews are obtained through the database of international journal providers such as Proquest, Google Scholar and Science Direct. Journals are taken from 2019 to 2021. Researchers used keywords namely transcultural nursing, breastfeeding (according to MESH / Medical Subject Heading) and selected full text. There were 895 findings, then narrowed by monitoring duplication and found 391 findings. Then there are restrictions on articles that are less relevant and found 402 articles, then narrowed down again with restrictions on full text articles that can be accessed found as many as 102 articles. Based on the determination of inclusion criteria obtained 20 appropriate articles. The process of searching for articles in the journal, described in the PRISMA chart flow as follows:

**Figure 1.**
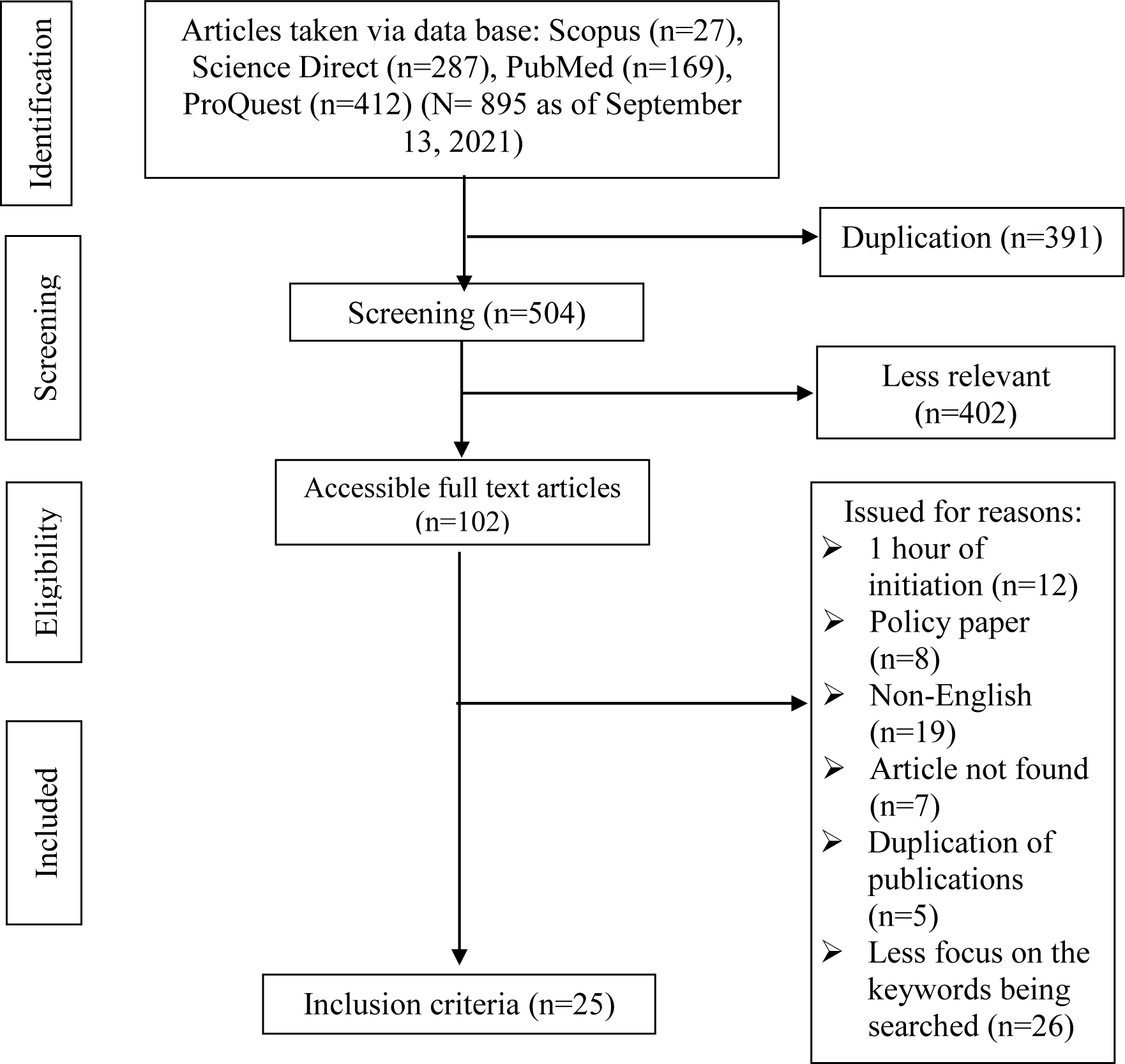
Flowchart of Prism

**Table 1.** Authenticity of Research

| RESEARCH TITLE | RESEARCH METHODS | RESEARCH RESULTS |
| --- | --- | --- |
| Wagner, S, et al. 2019. Breastfeeding Initiation and Duration in France: The Importance of Intergenerational and Previous Maternal Breastfeeding Experiences-Results from The Nationwide ELFE Study. *Midwifery*. Vol. 69. Pages: 67-75. | Design: Longitudinal Study<br>Subjects: breastfeeding mothers in Perancis number of 320 people<br>Variables: experience of breastfeeding mothers<br>Instrument: ELFE study<br>Analysis: chi square test | The effect of breastfeeding in infancy is very important especially for primipara mothers and in multipara mothers without prior breastfeeding experience. Found a link between initiation of breastfeeding and duration of breastfeeding in infants using ELFE scheme. |
| Tseng, JF, et al. 2020. Effectiveness of an Integrated Breastfeeding Education Program to Improve Self-Efficacy and Exclusive Breastfeeding Rate: A Single-blind, Randomised Controlled Study. *International Journal of Nursing Studies*. Vol. 111, November 2020, 103770. | Design: experimental (a single blind, randomized controlled trial with a parallel group design)<br>Subjects: primigravid mothers with gestational ages of 12-32 weeks at the prenatal clinic of the education hospital, and her partner selected by convenience sampling (N=104) and allocated to the intervention group and control group.<br>Variables: Breastfeeding education programs (free variables) and self-efficacy, infant feeding attitudes and breastfeeding practices (bound variables).<br>Instrument: Breastfeeding self-efficacy measured using Taiwan's version of self efficacy scale (BSES-SF) consists of 14 items to measure a breastfeeding mother's confidence. To assess the attitude of breastfeeding mothers, the Taiwan version of the Iowa Infant Feeding Attitude Scale (IIFAS) consisted of 17 items. To assess the level of breastfeeding used a self- | Intervention of breastfeeding education programs increases the mother's self-efficacy for breastfeeding, infant feeding attitudes and exclusive breastfeeding rates. |
|  | evaluation questionnaire with 3 assessment items: 1) exclusive, 2) Predominant and 3) formula milk. Analysis: independent t-tests, chi-square or Fisher's exact test using SAS software version 9.4 |  |
| Walsh, SM, et al. 2019. Effects of Early Initiation of Breastfeeding on Exclusive Breastfeeding Practices of Mothers in Rural Haiti. *Journal of Pediatric Health Care. Vo. 33. Number 5. September/Oktober 2019. Pages 561-567.* | Design: descriptive cross-sectional.<br>Subjects: mothers who had 195 breastfed babies collected data on 3 variable stages: early breastfeeding initiation (free variable) and exclusive breastfeeding (bound variable).<br>Instrument: check list<br>Analysis: bivariate analysis. | The initiation of early breastfeeding has an effect on the success of exclusive breastfeeding in rural Haiti so that EBI campaign activities to be launched especially by local leaders. Unique cultural influences are a determining factor in the success of this stage, but can also be an obstacle if the mother's beliefs are ignored. Promotive efforts are needed to convince every mother that the mother can breastfeed 6 months well. |
| Belachew, A. 2019. Timely Initiation of Breastfeeding and Associated Factors Among Mothers of Infants Age 0-6 Months Old in Bahir Dar City, Northwest, Ethiopia, 2017: A Community Based Cross-Sectional Study. *International Breastfeeding Journal. Vol. 14. No.5. Pages: 1-6.* | Design: cross sectional study<br>Subjects: 472 mothers who had babies less than 6 months of age in Bahir Dar City<br>Variables: breastfeeding initiation rate (bound variable) factors from mothers related to breastfeeding (free variables)<br>Instruments: questionnaire<br>Analysis: descriptive statistics and logistic regression | The implementation of breastfeeding initiation in Bahir Dar City is still less than optimal, mothers who get information about early breastfeeding initiation before can carry out EBI well. Other factors such as abdominal labor and lack of maternal knowledge, impact the implementation of EBI. |
| Mishbahatul, E, et al. 2018. Early Weaning Food for Infants (0-6 Months Old) in Madurese People Based on transcultural Nursing Theory. *IOP Publishing: 3<sup>rd</sup> International Conference on* | Design: cross sectional<br>Subjects: 61 mothers who had babies aged 0-6 months who had been given supplemental meals<br>Variables: Education, economics, politics and law, | Based on the transcultural nursing theory it shows a diversity of positive and negative values that affect early feeding to infants aged 0-6 months in Madura. |
| *Tropical and Coastal Region Eco Development.* | cultural, lifestyle, kinship and social values, religion and philosophy and technology (free variables) that affect early feeding in infants aged 0-6 months (bound variables)<br>Instruments: structured interview<br>Analysis: frequency and percentage distribution |  |
| Ali, F, et al. 2020. Prevalence of and Factors Associated with Early Initiation of Breastfeeding Among Women with Children Aged <24 Months in Kilimanjaro Region, Northern Tanzania: A Community Based Cross Sectional Study. *International Breastfeeding Journal*. Vol. 15. No. 80. Pages: 1-10. | Design: cross sectional<br>Subjects: 1644 mothers who had children aged <24 months in kilimanjaro region, Northern Tanzania<br>Variables: pre lacteal feeding, family support and health workers, maternal knowledge (free variables). Initiation of early breastfeeding (bound variable)<br>Instrument: questionnaire<br>Analysis: linear regression | It was found that the association of family support factors, health support, pre lacteal maanan and maternal awareness was associated with low EBI implementation in the Kilimanjaro region of Northern Tanzania.. |
| Abdulahi, M, et al. 2021. Breastfeeding Education and Support to Improve Early Initiation and Exclusive Breastfeeding Practices and Infant Growth: A Cluster Randomized Cotrolled Trial from a Rural Ethiopian Setting. *Nutrients*. Vol. 13. No. 1204. Pages: 1-15. | Design: experiments<br>Subject: pregnant women trimester III to 5 months post-delivery (intervention group: 249 people and control group: 219 people)<br>Variables: Education and breastfeeding support (free variables), initiation of early breastfeeding, breastfeeding practices and infant growth (bound variables)<br>Instruments: instructed interviews and questionnaire<br>Analysis: linear regression | There is an influence of breastfeeding education and support on early breastfeeding initiation, breastfeeding practices and fetal growth.. |
| Namasivayam, V, et al. 2021. Association of Prenatal Counselling and Immediate Postnatal Support with Early Initiation of Breastfeeding in Uttar Pradesh, India. *International* | Design: cross sectional study<br>Subject: mother with child born live 0-59 days in 25 districts of Uttar Pradesh, India, numbering 9124 people | There is a significant link between community-based prenatal counseling and postnatal support at the home with early breastfeeding initiation.. |
| *Breastfeeding Journal*. Vol. 16. No. 26. Pages: 1-11. | Variables: prenatal community-based counseling, postnatal support at the scene of delivery (free variable) and early breastfeeding initiation (bound variable)<br>Instrument: questionnaire<br>Analysis: logistic regression bivariate |  |
| Ralhana, S, et al. 2019. Early Initiation of Breastfeeding and Severe Illness in The Early Newborn Period: An Observational Study in Rural Bangladesh. *PLOS Medicine*. Pages: 1-17. | Design: experiments<br>Subject: mothers with newborns amounted to 30,646 from 2013-2015 who were distinguished in initiation groups in 1 hour, 1-24 hours, 24-48 hours, over 48 hours and not breastfed.<br>Variables: initiation of early breastfeeding in mothers with newborns (free variables)<br>Instrument: check list<br>Analysis: logistic regression | Initiation of early breastfeeding within an hour of birth can significantly decrease the incidence of the disease in newborns. |
| Woldeamanuel, BT. 2020. Trends and Factors Associated to Early Initiation of Breastfeeding, Exclusive Breastfeeding and Duration of Breastfeeding in Ethiopia: Evidence from The Ethiopia Demographic and Health Survey 2016. *International Breastfeeding Journal*. Vol. 15. No. 13. Pages: 1-13. | Design: cross sectional study<br>Subjects: mothers who have children aged 0-6 months in Ethiopia numbering 5,122 people<br>Variables: child sex residence, birth type, birth weight of baby and family type (free variable), initiation of early breastfeeding, exclusive breastfeeding duration, duration of breastfeeding (bound variable)<br>Instrument: check list<br>Analysis: multivariate logistic regression | Rural residences, female gender, home labor, cesarean delivery, small birth weight babies and large family sizes are associated with late initiation of breastfeeding.. |
| Shofiya, D, et al. 2020. Nutritional Status, Family Income and Early Breastfeeding Initiation as Determinants to Successful | Design: cross sectional study<br>Subjects: mothers who have babies aged 6-24 months a total of 273 people | Nutritional status before pregnancy tends to benefit family income, therefore starting early breast milk needs to be improved for |
| Exclusive Breastfeeding. *Journal of Pulic Health Research*. Vol. 9, No. 1814. Pages:110-113. | Variables: maternal age, Education, family income, frequency of pregnancy examinations, nutritional status before pregnancy, place and manner of delivery, gestational age at childbirth (free variable), initiation of early breastfeeding and exclusive breast milk (bound variable)<br>Instrument: check lyst<br>Analysis: linear regression | the success of exclusive breastfeeding. |
| Nielsen, C, et al. 2021. Breastfeeding Initiation and Duration After High Exposure to Perfluoroalkyl Substances (PFAS) Through Contaminated Drinking Water: A Cohort Study from Ronneby, Sweden. *Environmental Research*. Pages: 1-9 | Design: cross sectional study<br>Subjects: infants born in 1999-2009 numbering 2,374 people<br>Variables: exposure to PFAS (free variable), initiation and duration of breastfeeding (bound variables)<br>Instrument: questionnaire<br>Analysis: chi square test | There is a relationship between PFAS exposure and initiation and duration of breastfeeding. |
| Yunitasari, E, et al. 2020. Factors Associated with Exclusive Breastfeeding Based on Transcultural Nursing. *EurAsian Journal of BioSciences*. Vol. 14. Pages: 2757-2766. | Design: descriptive analytics with cross sectional approach<br>Subjects: mothers who have babies aged 6-12 months amounting to 289 people<br>Variables: technological factors, religious and philosophical factors, social factors, cultural values and way of life, political and legal factors, economic factors and educational factors (free variables), exclusive breastfeeding based on transcultural nursing (bound variables).<br>Instrument: questionnaire<br>Analysis: chi square test | There is a relationship of technological, religious and philosophical factors, socio-cultural values and ways of life, politics and law, economics and education to exclusive breastfeeding based on transcultural nursing.. |
| Karnasih, IGA. 2021. Effectiveness of Leininger's Transcultral-based GATHER Counseling | Design: post test experiment only control group design<br>Subject: 308 pregnant | Mothers who were given counseling model GATHER based on |
| Model on Exclusive Breastfeeding. *Health Nations*. Vol. 5. No. 5. Pages: 156-162. | women trimester III divided into 2 groups<br>Variables: GATHER counseling model (free variable), exclusive breastfeeding (bound variable)<br>Instruments: questionnaire and observation<br>Analysis: Mann Whitney-U test | transcultural nursing more who breastfeed exclusively. |
| Azza, A, et al. 2017. The Application of Transcultural Nursing Model in Perspective of Madura Culture Improving Breastfeeding Mother's Behavior in Jember. *Jurnal INJEC*. Vol. 2. No. 1. Pages: 9-16. | Design: quasi experiment post test design with control group<br>Subjects: breastfeeding mothers who have babies aged 1-6 months as much as 50 samples divided into 2 groups<br>Variables: model nursing transcultural nursing (free variable), breastfeeding (bound variable)<br>Instrument: questionnaire<br>Analysis: paired sample t test | The application of cultural modification is able to improve the behavior of breastfeeding mothers. |
| Kuswara, K, et al. 2021. Breastfeeding and Emerging Motherhood Identity: An Interpretative Phenomwnological Analysis on First Time Chinese Australian Mothers' Breastfeeding Experiences. *Women and Birth*. Vol. 34. Pages: 292-301. | Design: interpretative analysis<br>Subjects: Australian Chinese mothers numbering 11 people who are breastfeeding<br>Variables: first-time breastfeeding experience in mothers of Chinese Australian descent (free variables)<br>Instrument: structured interview<br>Analysis: descriptive statistics | Breastfeeding becomes an enjoyable experience for mothers of Chinese Australian descent because mothers feel increasingly confident and a sense of motherhood that increases motivation to breastfeed. |
| Homayi, SG, et al. 2020. Skin to Skin Contact, Early Initiation of Breastfeeding and Childbirth Experience in First Time Mothers: A Cross Sectional Study. *Journal of* | Design: cross sectional study<br>Subjects: primipara mothers who give birth per vaginam number of 800 people<br>Variables: skin-to-skin contact (free variables) and | There is a link between skin contact and early breastfeeding initiation within the first hour of giving birth. |
| *Neonatal Nursing*. Vol. 26. Pages: 115-119. | initiation of early breastfeeding within the first hour of childbirth (bound variable)<br>Instruments: questionnaire and check list<br>Analysis: multivariable logistic regression test |  |
| Alghamdi, S, et al. 2017. Racial and Ethnic Differences in Breastfeeding, Maternal Knowledge, and Self Efficacy Among Low-Income Mothers. *Applied Nursing Research*. Vol. 37. Pages: 24-27. | Design: randomized controlled trial experiment.<br>Subjects: low-income mothers who have babies aged 0-6 months amounting to 540 people<br>Variables: maternal knowledge and self-efficacy (free variables), feeding and tendencies in breastfeeding (bound variables)<br>Instrument: questionnaire<br>Analysis: chi square test | There is no significant correlation between maternal knowledge, self-efficacy in feeding the baby and the tendency to breastfeed.. |
| Walsh, SM, et al. 2019. Effects of Early Initiation of Breastfeeding on Exclusive Practices of Mothers in Rural Haiti. *Journal of Pediatric Health Care*. Vol. 33. No. 5. Pages: 561-567. | Design: cross sectional study<br>Subject: postpartum mothers in rural Haiti number 195 people<br>Variables: early breastfeeding initiation (free variable), exclusive breastfeeding (bound variable)<br>Instrument: questionnaire<br>Analysis: chi square test | There is an influence of early breastfeeding initiation on breastfeeding exclusively. |
| Potts, KS, et al. 2021. Early Breastfeeding and Complementary Feeding in Ethiopia: Cross Sectional Data From Implementation of Nutrition Programming in Regional Inequalities. *Heliyon*. Vol. 7. Pages: 1-9 | Design: cross sectional study<br>Subjects: mothers who have babies aged 0-36 months amounting to 1,299 people<br>Variables: implementation of nutrition programs (free variables), initiation of early breastfeeding and feeding (bound variables)<br>Instruments: questionnaire<br>Analysis: chi square test | There was a link between early breastfeeding and end-of-breast milk feeding practices with certain socio-demographic characteristics.. |
| Wen, J, et al. 2021. Effects of a Theory of Planned Behavior based Intervention | Design: randomized controlled trial Experiment<br>Subjects: mothers who gave | Mothers who were given more behavioral interventions who provided |
| on Breastfeeding Behaviors After Cesarean Section: A Randomized Controlled Trial. *International Journal of Nursing Sciences*. Vol. 8. Pages: 152-160. | birth sc number of 32 people<br>Variables: intervention-based behavioral theory (free variables) and breastfeeding behavior (bound variables)<br>Instrument: questionnaire<br>Analysis: chi square test | exclusive breast milk than mothers who were not given interventions |
| O'Relly, SL, et al. 2021. Latch On: A Protocol for a Multi Centre, Randomised Controlled Trial of Perinatal Support to Improve Breastfeeding Outcomes in Women with a Raised BMI. *Contemporary Clinical Trials Communications*. Vol. 22. Pages: 1-6. | Design: randomized and controlled experiment<br>Subjects: obese mothers who breastfed babies 0-3 months totaling 220 people<br>Variables: multifactor interventions (free variables) and breastfeeding behavior (bound variables)<br>Instruments: questionnaire<br>Analysis: logistic regression | Multifactor support affects breastfeeding success in obese mothers |
| Padashian, F, et al. 2021. Examining Exclusive Breastfeeding in Iranian Mothers Using The Five Factor Model of Personality Traits. *Journal of Taibah University Medical Sciences*. Pages: 1-6. | Design: cross sectional study<br>Subjects: mothers who have babies aged 6-12 months numbering 120 people<br>Variables: five models of maternal personality (free variables), exclusive breastfeeding (bound variables)<br>Instruments: questionnaire<br>Analysis: Pearson correlation and multiple regression | A mother's personality has the potential to influence the success of exclusive breastfeeding and can be a tool to reduce barriers in exclusive breastfeeding. |
| Forster, DA, et al. 2019. Proactive Peer (Mother to Mother) Breastfeeding Support by Telephone (Ringing up About Breastfeeding Early / RUBY): A Multicentre, Unblinded, Randomised Controlled Trial. *EClinicalMedicine*. Vol. 8. Pages: 20-28. | Design: randomized controlled experiment<br>Subjects: primipara mothers who are breastfeeding babies 0-6 months a number variables: proactive telephone-based peer support during the postnatal period (free variable), proportion of breastfed infants at six months of age (bound variable)<br>Instrument: questionnaire<br>Analysis: descriptive statistics | Proactive phone-based peer support during the postnatal period had an impact on an increase in the proportion of breastfed babies up to six months of age. |

## RESULTS AND DISCUSSION

Early Breastfeeding Initiation (EBI) is a process of allowing the baby with his own instinct to breastfeed immediately within the first hour after birth, along with contact between the baby’s skin and the mother’s skin, until the baby suckles himself (Ministry of Health, 2008). The way babies do EBI is called The Best Crawl or crawling looking for breasts (Maryunani, 2012). In foreign terms EBI is called early initiation breastfeeding. When a healthy baby is placed on the mother’s stomach or chest immediately after birth and skin to skin contact is an amazing show, the baby will react by the stimulation of the mother’s touch, the baby will move over the mother’s abdomen to look for the mother’s breasts (Roesli, 2012). The many benefits obtained by babies through the EBI process, causing the World Health Organization (WHO) to recommend EBI be implemented as part of the labor process, and in Indonesia EBI has begun to be socialized since August 2007 (Kemenkes RI, 2012).

Pitaloka research (2017) in Yunitasari, E, et al (2021) states that public figures and religious figures influence the behavior of mothers in breastfeeding their babies. People believe the tradition and culture of combining milk with water or additional food will accelerate the growth of the baby. While Karnasih’s research, IGA (2021) mentions one of the factors postpartum mothers in East Java do not breastfeed their babies is due to socio-cultural related factors adopted by mothers and their families. Mishbahatul (2017) in his research conducted in Madura, found the fact that the practice of breastfeeding companion feeding before the baby is 6 months old because of the belief that babies given breast milk companion food earlier will grow better and sleep better. Based on some of these researches, culture and tradition are one of the factors that affect breastfeeding in infants in Indonesia.

Early breastfeeding initiation policy has been socialized in Indonesia since August 2007 (Roesli, 2012). The World Health Organization (WHO) has recommended to all infants to get colostrum on the first and second day to fight various infections and get exclusive breast milk for 6 months (Kemenkes, 2012).

Research conducted by Susanto, T, et al. (2021) in Jember Regency, East Java found that many mothers do not practice early breastfeeding initiation and exclusive breastfeeding due to ignorance, unwillingness and incompetence that are influenced by the lack of family support. Habits in the family that give up the entire business of breastfeeding to the mother without optimal support, turned out to be an obstacle to the success of breastfeeding in the mother. Mishbahatul, E, et al. (2019) revealed the tradition and belief in families in Madura that the earlier giving solid food to babies will accelerate the baby’s weight gain, this belief is certainly not in line with the theory regarding the practice of early breastfeeding initiation.

As research conducted by Akter, F, et al. (2019); Bolton, KA, et al. (2019); Camier, A, et al. (2021); Dowle, KM, et al (2021) the immigrant population is actually more likely to initiate early breastfeeding that continues in exclusive breastfeeding compared to natives in the United States because of the belief that milk is rich in benefits when compared to other types of food for infants. While the study conducted by Mavisoa, MK, et al. (2021) in Papua New Guinea, not all primipara mothers can practice early breastfeeding initiation even though this program has been implemented as part of the labor process. This is due to the imbalance of gender roles as a cultural heritage in Papua New Guinea, where fathers play an important role in decision-making in the family. While research conducted by Aakre, I, et al. (2019); Goes, FGB, et al. (2019); Cotelo, MdCS, et al. (2019); Yugistyowati, A, et al. (2020) in Kulon Progo Regency actually found a cultural practice that supports the practice of breastfeeding in infants. Mothers and families believe that the more often the baby is breastfed, the baby will grow well and rarely get sick.

Research conducted by Abuhammad, S, et al. (2021) in Jordan found that the culture of mother and baby attachment during childbirth helps increase the production of oxytocin that can affect milk production, so health workers are advised to pay attention to this aspect in the provision of care to the mother by doing follow-up to the mother and baby after birth. While research conducted by Potts, KS, et al. (2021) on breastfeeding mothers in Ethiopia found a cultural relationship of eating in nursing mothers that can affect the nutritional status of infants, it is necessary to approach socio-culturally to change eating habits in the family so that the nutritional needs of babies breastfed by their mothers can be met through the adequacy of feeding in the mother.

Cultural aspects that are inherited for generations in the family, need to get special attention in providing nursing care to post partum mothers. Karnasih, IGA. (2021) through her research found that the application of a culture-based nursing model (Leininger) can help increase maternal knowledge about the importance of early breastfeeding initiation practices, because mothers have the full support of the family and cultural aspects brought by mothers and families are not used as obstacles in breastfeeding practices, but instead be used as boosters of maternal and family beliefs so that breastfeeding practices can be done exclusively.

This research is also in line with research conducted by Edwards, R, et al. (2021); Naragale, YM, et al. (2020); Naeini, MA, et al. (2021); Yunitasari, E, et al. (2020) who found that a culture-based approach turned out to be very good for mothers and families to help mothers carry out optimal breastfeeding practices, because the beliefs that exist in mothers are strengthened with the full support of the family.

## CONCLUSION

As a government commitment to EBI practices, the Indonesian government supports WHO and UNICEF policies recommending EBI as a life-saving measure, as EBI can save 22% of babies who die before the age of one month. Breastfeeding for the first hour of life that begins with skin contact between mother and baby is declared a global indicator. EBI is a government program so that it is expected that all health workers at all levels of health services can socialize and implement the program so that it is expected that quality Indonesian human resources will be achieved.

As a nurse, in providing care to mothers during breastfeeding, the ability to recognize the cultural perspective that the mother believes to be a tradition that is believed to be true in the practice of breastfeeding needs attention. By knowing the cultural perspective that the mother believes in the practice of breastfeeding, nurses can use culture-based communication strategies to mediate these beliefs so that mothers and families do not feel disturbed in the act of denial to tradition, but the practice of breastfeeding can be implemented in accordance with the recommendations that have been set.

## Data Availability

All data produced in the present work are contained in the manuscript

